# Association of pre-pandemic respiratory system diseases with Long COVID: a population-based case-control study

**DOI:** 10.1101/2024.06.07.24308594

**Authors:** Pia Lindberg, Sebastian Lindblom, Gunnar Ljunggren, Seika Lee, Iryna Kolosenko, Michael Runold, Artur Fedorowski, Caroline Wachtler, Kristina Piontkovskaya, Axel C. Carlsson, Åsa M. Wheelock

## Abstract

**Objectives:** Long COVID, defined as diverse symptoms persisting >3 months post-infection, remains a major post-pandemic healthcare burden. Here we investigate risk factor posed by pre-existing respiratory symptoms and illnesses for development of long COVID, with focus on individuals with mild-to-moderate COVID-19 at the primary infection, that did not require hospitalization during the primary SARS-CoV-2 infection.

**Methods:** This case-control study was designed to investigate the prevalence of respiratory system-related diagnoses in adult, non-hospitalized long COVID patients (cases) compared to matched controls without a history of long COVID. Data was extracted from the Stockholm Region’s database (VAL) and included diagnoses 12 months pre- and 6 months post-long COVID diagnosis as well as pre-pandemic diagnoses (year 2019). Adjusted logistic regression models were applied.

**Results:** Patients with Long COVID displayed higher frequencies of pre-pandemic respiratory conditions (year 2019) as well as 12 months before long COVID diagnosis compared to controls, including acute upper respiratory tract infections (men: Odds ratio (OR) 2.47, women: OR 2.22), asthma (men: OR 1.76, women: OR 1.95), and bronchitis (men: OR 2.15, women: OR 2.71). ORs for asthma were the highest 12 months before long COVID diagnosis (men: OR 4.18, women: OR 3.76).

**Conclusion:** Patients with Long COVID with a mild-to-moderate primary SARS-CoV-2 infection had higher prevalence of pre-existing respiratory conditions than controls, suggesting that respiratory diseases including asthma were a significant risk factor for long COVID also in the non-hospitalized population. Understanding the link between chronic respiratory illnesses and long COVID is vital for refining clinical strategies and improving outcomes in post-viral conditions.

**Key take-home message:** Pre-pandemic respiratory diagnoses, including asthma, as well as female sex represent significant risk factors for developing long COVID in individuals with a mild-to-moderate primary SARS-CoV-2 infection not requiring hositalization.

## INTRODUCTION

Long COVID also referred to as post-acute Sars-CoV-2 syndrome (PACS) or post-COVID [1–5] is a new diagnosis with a spectrum of diverse persistent symptoms involving multiple organ systems. Long COVID is defined by symptoms associated with SARS-Cov-2 infection such as chronic cough, chest pain, fatigue, brain fog, dizziness, gastrointestinal symptoms, palpitations, changes in libido and sexual capacity, loss or change in the perception of smell or taste persisting 3 months after the primary infection. In autumn 2020, WHO introduced the first definition and a diagnostic code for post-COVID-19 condition [6]. In 2021, a Delphi process was conducted to provide consensus guidelines for diagnosis of the new post-COVID-19 condition [7, 8]. This consensus specified that symptoms of the syndrome can be diverse and persist for at least three months, can fluctuate in intensity over time and cannot be attributed to any other diagnosis. The risk factors and mechanisms associated with long COVID remain unclear, and it is unlikely that a single factor accounts for the wide range of symptoms affecting different organ systems. At this point it is evident that long COVID is an umbrella diagnosis composed of multiple sub phenotypes, each influenced by distinct risk factors, biological mechanisms, and disease courses. Various elements, such as genetics, age, gender, pre-existing health conditions, microbiome composition, and viral traits, may trigger different pathological responses [9].

Similarities between long COVID and post-intensive care syndrome (PICS) symptoms, and the higher prevalence of long COVID in individuals who received intensive care for severe COVID-19 during the acute infection initially led to the theory that these symptoms primarily represented sequelae of intensive care, common in other conditions [10]. However, a range of studies have indicated a wide variation in prevalence for long COVID. Some studies show long COVID prevalence around 10% among non-hospitalized patients with a mild infection [1, 3]. Longitudinal studies suggest that up to 46% of patients have persistent symptoms 12 months after a non-severe COVID-19 infection [11, 12]. A review by Davies et al. noted that long COVID affects individuals across all age groups and regardless of the severity of the acute phase [4]. However, the highest incidence of long COVID is seen in those between 36 and 50 years of age, with female predominance, with the majority of cases occurring in non-hospitalized patients who had an asymptomatic or mild-to-moderate initial SARS-COV-2 infection (from here referred to as the non-hospitalized long COVID population) [4].

In this study, we aimed to investigate the association between pre-pandemic respiratory illnesses and long COVID prevalence in the non-hospitalized long COVID population, with a focus on sex differences and asthma prevalence.

## METHODS

### Study Database

The Stockholm County Healthcare Region (Stockholm Region) provides healthcare services under the governance of the Stockholm Region. With approximately 2.5 million residents, the Stockholm Region is the largest in Sweden, covering a variety of urban and rural areas. Healthcare is meticulously documented in the Stockholm Regional Health Care Data Warehouse (VAL), with a documented credibility through contribution to national health registers and benchmarking reports [13]. Since 1997, diagnoses have been systematically coded according to the World Health Organization’s International Classification of Diseases, 10^th^ revision (ICD-10), ensuring standardization and consistency in healthcare data management [6]. The authors had access to diagnoses recorded at all care levels in the Region, including both primary and specialist care.

However, as with any registry-based study, some missing data cannot be ruled out. In addition to registry data, relevant literature on post-COVID respiratory conditions was identified using the CORACLE platform [14].

### Study Population

The MIRACLE-S (Multimorbidity Integrated Registry Across Care Levels in Stockholm) study cohort comprised adults 18 years of age and older residing in the Stockholm Region for the entire duration of the study, January 2019 to February 2022 [15]. Individuals who relocated in or out of the region or died during this period were excluded. The inclusion criteria for long COVID patients (cases) were post-COVID-19 diagnosis U09.9 (ICD-10) obtained for the first time between January 1 and December 31, 2021, in primary or specialized healthcare settings. Individuals who were hospitalized during the acute phase of COVID-19 were excluded. Each case was matched with 10 controls based on age, sex and neighborhood socioeconomic status. Based on the date when cases obtained their long COVID diagnosis, diagnostic codes for symptoms and conditions related to post-COVID with respiratory involvement were extracted (3,4) from three different timepoints: diagnoses recorded during 2019, 12 months prior to long COVID diagnosis, as well as 6 months after the date of U09.9 diagnosis for both cases and matched controls. Since access to data from the registry was limited until February 10, 2022, the 6-month post-PACS diagnostic timepoint was shorter for some study subjects (minimum of 40 days). The study design is illustrated in Figure 1.

**Figure 1.**
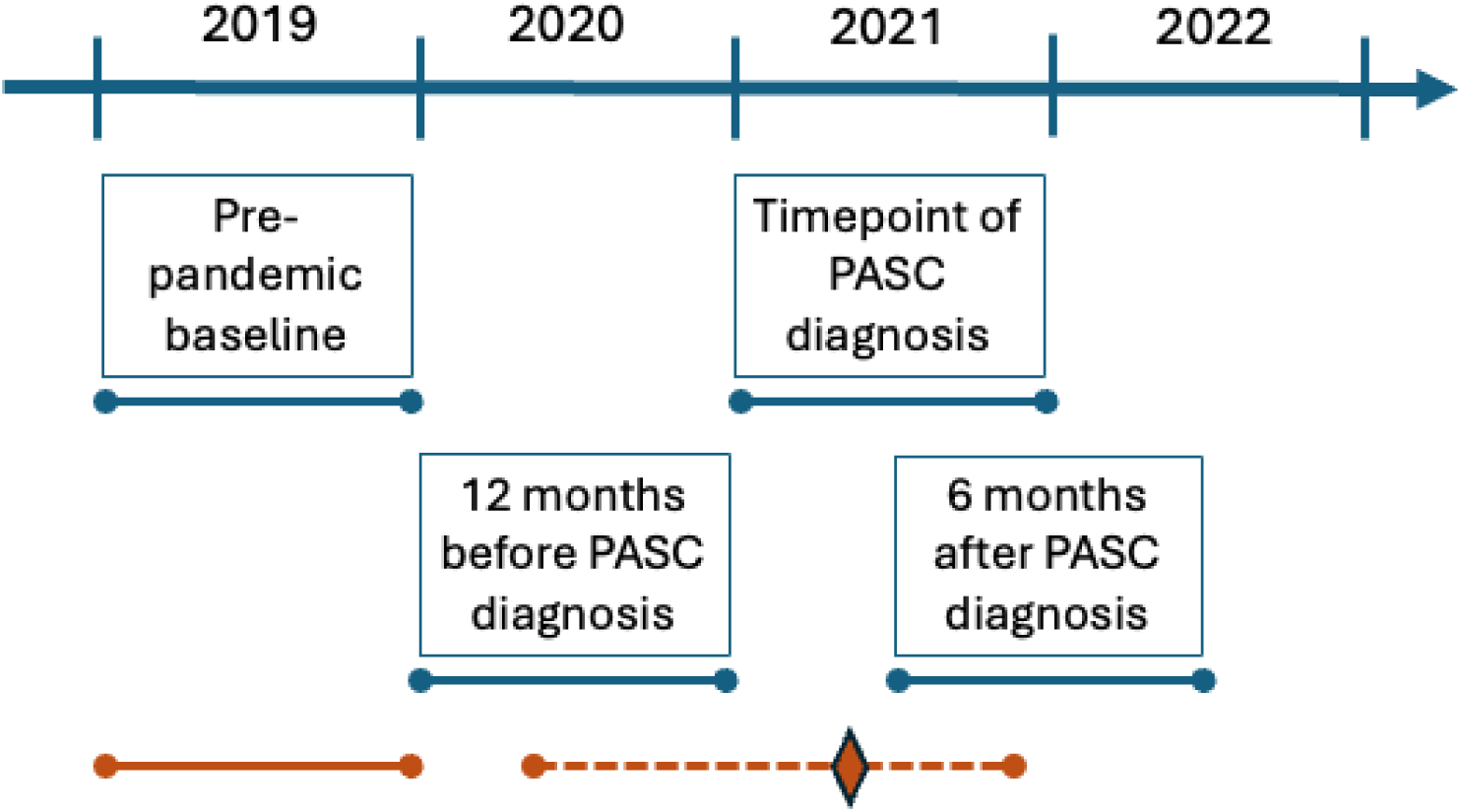
Outline of the study design for the data collection time points. Patients diagnosed with long COVID during 2021 (n= 5 589) were matched 1:10 with non-long COVID controls (n= 47 561) based on age, sex, and socioeconomical neighborhood on the corresponding date. The timepoint of data collection is outlined for the full study in teal, as well as exemplified for one individual in orange (diamond: date of long COVID diagnosis). Data on respiratory diagnoses were retrieved retrospectively for the Timepoint of long COVID diagnosis, 12 months before long COVID diagnosis, for the pre-pandemic year of 2019, as well as 6 months after long COVID diagnosis.

### Diagnoses

ICD-10 diagnoses applied in this study are presented in Supplementary Table 1 [6]. In brief, the diagnosis code for long COVID is U09.9, post COVID-19, and the respiratory diagnoses investigated include acute upper respiratory tract infection (J06.9), acute bronchitis (J20.9, J40.9), asthma (J45.1, J45.8, J45.9, J46.9), cough (R05), dyspnea (R06.0) and other and unspecified respiratory disorders (R06.8).

### Neighborhood socioeconomic status

The Mosaic tool was utilized to categorize various municipalities and regions within the area based on socioeconomic status. Originally designed to optimize sales efficiency, Mosaic has demonstrated credibility in epidemiological studies due to its ability to generate multivariable models, encompassing over 400 variables. Through the utilization of Mosaic, three levels of socioeconomic status—high, medium, and low—were delineated [16, 17].

### Ethical approval

Ethical approval was obtained from the Swedish Ethical Review Authority (Case no. 2021-01016 and 2021-05735).

### Statistical methods

Conditional logistic regression was used to calculate odds ratios (OR) with 99% confidence intervals (CI) for the occurrence of defined diagnoses in cases versus controls. Statistical analysis and data management were conducted using SAS software, version 9.4 (SAS Institute Inc., Cary, NC, USA).

## RESULTS

We identified 5589 individuals (69.1% females), mean age 47 years (47-48) who met the inclusion criteria for cases. These individuals were matched with 47,561 controls.

Demographic details are presented in Supplementary Table 2.

The odds ratios for the prevalence of the investigated respiratory symptoms and diagnoses between long COVID patients and matched controls for year 2019 (pre-pandemic), 12 months before long COVID diagnosis, and 6 months after long COVID diagnosis are shown in Figure 2 for both sexes, and in Table 1 and Table 2 for females and males respectively.

**Figure 2.**
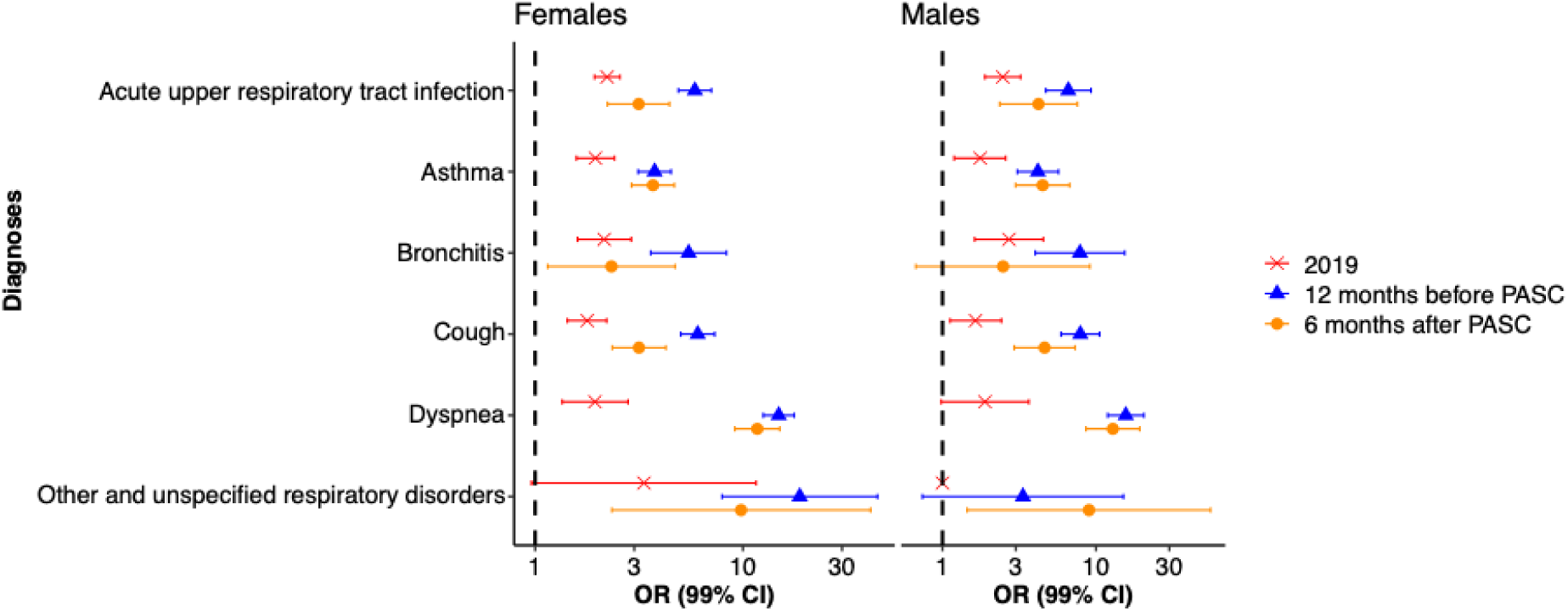
Summary of the association between investigated respiratory symptoms and diagnoses in relation to long COVID. The odds ratios (OR) and 99% CI are presented for females and males respectively in 2019, 12 months prior to long COVID diagnosis, and 6 months after long COVID diagnosis.

**Table 1.** Association between long COVID and respiratory system diagnoses in women shown as odds ratios in the year 2019, 12 months before long COVID diagnosis, and 6 months after the long COVID diagnosis. Female Cases: n=3862; Female Controls: n=32151.

| Diagnosis | Pre-pandemic<br>(year 2019) | 12 months pre-<br>long COVID<br>diagnosis | 6 months post-long COVID<br>diagnosis* |
| --- | --- | --- | --- |
| **Other and unspecified**<br>**respiratory disorders R06.8** | 3.34<br>(0.96-11.6) | 18.8<br>(7.94-44.4) | 9.80<br>(2.34-41.1) |
| **Acute upper respiratory**<br>**tract infection J06.9** | 2.22<br>(1.93-2.56) | 5.87<br>(4.89-7.05) | 3.15<br>(2.23-4.43) |
| **Bronchitis J20.9, J40.9** | 2.15<br>(1.60-2.90) | 5.48<br>(3.60-8.33) | 2.33<br>(1.15-4.72) |
| **Asthma J45.1, J45.8, J45.9,**<br>**J46.9** | 1.95<br>(1.58-2.41) | 3.76<br>(3.13-4.51) | 3.69<br>(2.91-4.68) |
| **Dyspnea R06.0** | 1.94<br>(1.35-2.80) | 14.9<br>(12.5-17.6) | 11.7<br>(9.14-15.0) |
| **Cough R05** | 1.78<br>(1.43-2.21) | 6.06<br>(5.02-7.31) | 3.16<br>(2.35-4.26) |

\*The 6-month time point was truncated for some individuals due to data access. Please see methods.

**Table 2.**
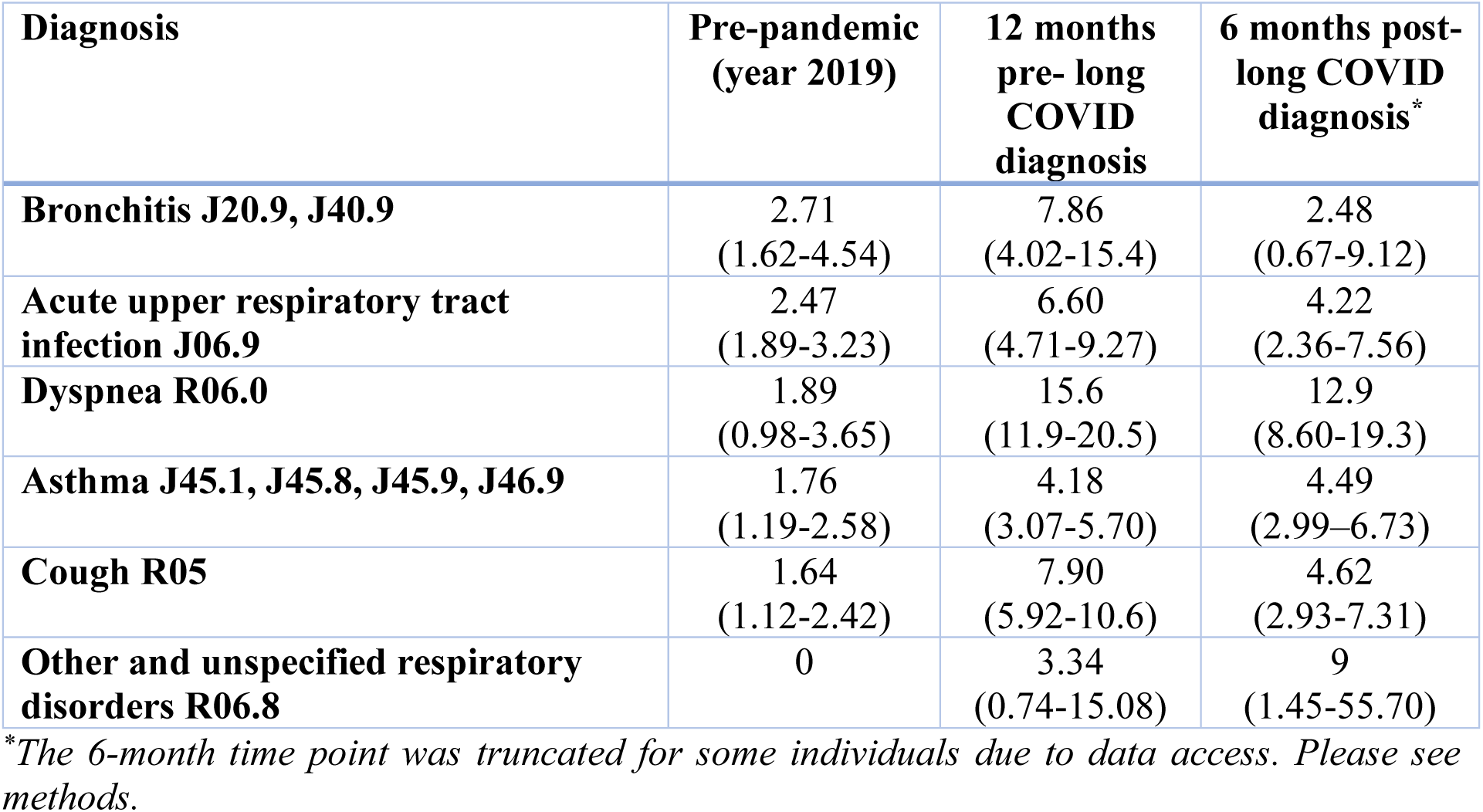
Association between long COVID and respiratory system diagnoses in men shown as OR and 99% CI in the year 2019, 12 months before the long COVID diagnosis, and 6 months after the long COVID diagnosis. Male Cases: n=1727; Male Controls: n=15410.

There was a higher prevalence of all studied diagnoses in long COVID patients compared with their matched controls both prior to and after the COVID-19 pandemic (Figure 2).

Diagnoses with the highest odds ratio before the pandemic (2019) were acute upper respiratory tract infection (J06.9) and bronchitis (J20.9, J40.9) for both sexes, as well as dyspnea (R06.0) for males (Figure 2, Table 1-2). Diagnoses with the highest odds ratio 12 month before long COVID were symptom diagnoses dyspnea (R06.0) and cough (R05) for both sexes, as well as unspecified respiratory disorders (R06.8) for females and bronchitis (J20.9, J40.9) for males. Please note that the 12-month pre-diagnosis time point precedes the existence of a long COVID diagnosis altogether or may represent a time point where the individual suffered from undiagnosed long COVID, why the 2019 time point represents a mor accurate reference point for pre-existing risk factors for long COVID.

In terms of asthma diagnosis (J45), we obtained OR=1.95 for females and OR=1.76 for males in 2019, 3.76 for females and 4.18 for males 12 months before long COVID diagnosis, and 3.69 for females and 4.49 for males 6 months after long COVID diagnosis.

The diagnoses with the highest OR six months after long COVID diagnosis were dyspnea (R06.0) and other and unspecified respiratory disorders (R06.8) for both sexes, as well as asthma (J45.1, J45.8, J45.9, J46.9) for females and cough (R05) for males (Figure 2, Table 1-2).

Most of the diagnoses exhibited significantly higher odds ratios, both among females and males 12 months prior to the long COVID diagnosis. Importantly, the odds ratios 6 months after long COVID diagnosis were in general lower compared with the period prior to long COVID diagnosis (Figure 2, Table 1-2), with the exception of asthma and dyspnea which remained at elevated levels in the long COVID groups of both sexes. This may represent an effect of the correct diagnosis of long COVID having been defined at this time point.

## DISCUSSION

In this study we investigated the prevalence of pre-pandemic respiratory system diagnoses and related respiratory symptoms as a predisposing risk factor for long COVID. Focus was placed on individuals who did not require hospitalization during their primary SARS-CoV-2 infection who received a long COVID diagnosis in Region Stockholm during 2021, thereby capturing primarily long COVID associated with the wt and alfa SARS-CoV-2 variants. Overall, the long COVID group displayed a higher frequency of respiratory illnesses and symptom diagnoses prior to the primary SARS-CoV-2 infection compared to matched controls: In the pre-pandemic year of 2019, individuals that later developed long COVID had a higher prevalence of acute upper respiratory tract infection, asthma, and bronchitis compared to their matched non-long COVID controls. The higher pre-pandemic prevalence was observed in both men and women.

More than two thirds of patients diagnosed with long COVID during 2021 in the Stockholm region were females, thereby corroborating previous findings that long COVID is more prevalent among females [18–22], including non-hospitalized cases. Furthermore, our study highlights that individuals who developed long COVID following a mild-to-moderate SARS-CoV-2 infection had a higher prevalence of various respiratory symptoms and asthma, particularly non-allergic asthma, than their matched controls before to the COVID-19 pandemic. We found significantly higher odds ratios of asthma among long COVID cases compared to controls, both before and after long COVID diagnosis (Table 1-2). Female sex and asthma have been suggested as risk factors for long COVID in prior studies. Jacobs et al. reported an adjusted odds ratio of 1.99 for female sex being associated with persistent symptoms after COVID-19, whereas O’Keefe and colleagues found that individuals with asthma had an increased risk of developing long COVID with an odds ratio of 1.54 [23, 24]. Other work shows similar associations regardless of hospitalization status [21]. Moreover, comorbid conditions such as asthma and a history of healthcare usage were found to be more strongly associated with long COVID development in non-hospitalized individuals than in those who had been hospitalized for severe COVID-19 [21]. A recently published nationwide observational study utilizing Swedish registries with matched controls also identified the female sex as a risk factor for being diagnosed with long COVID [25]. This study included all individuals with a registered COVID-19 diagnosis, encompassing both hospitalized and non-hospitalized cases [25]. However, limited access to nationwide primary care data may have restricted certain diagnoses. In addition to female sex, the highest risk factors associated with long COVID were ICD-10 symptoms and diagnostic codes [6] were dyspnea, abnormal findings on pulmonary diagnostic imaging, and chest pain [25]. Our study showed similar findings also in individuals that developed long COVID after a mild-to-moderate primary SARS-CoV-2 infection, not requiring hospitalization.

Taken together, our data show that patients with long COVID related to mild-to-moderate primary infections have higher proportions of pre-pandemic respiratory symptoms and illnesses than their controls. Residual predisposition to acute and chronic inflammatory diseases represents putative mechanism for developing both respiratory illnesses and long COVID. Prospective data revealed that baseline respiratory symptoms strongly predicted persistent post-COVID manifestations in outpatient populations, aligning with your observed pre-pandemic risk patterns [26]. SARS-CoV-2 is a pantropic virus, and pre-infection respiratory conditions may facilitate viral invasion and replication, promote viral persistence and inhibit virus elimination promoting post-viral complications. Further, an impaired immune response to respiratory pathogens that are associated with chronic conditions such as asthma and chronic bronchitis may be also a risk factor for long COVID [27]. Finally, the cumulative effect of prior infections and inflammatory conditions in the respiratory tract might trigger the long COVID cascade, such as is known for asthma. Our observations are in agreement with a study by Al-Aly et al. demonstrating that individuals with prior respiratory conditions such as asthma and chronic bronchitis are more likely to develop long COVID [9]. This supports the hypothesis that prior respiratory illnesses, such as asthma and chronic bronchitis, may predispose individuals to the development of long COVID, potentially through an increased susceptibility to post-viral complications and chronic inflammation. Liew et al. recently identified aberrant immune activation, particularly of the IFN-γ/IL-6 pathways, as potential mechanisms linking acute infection to long-term multisystem involvement, suggesting that pre-existing respiratory conditions sharing overlapping inflammatory pathways may predispose to long COVID [28].

The higher pre-pandemic incidence of respiratory illnesses and diagnoses in individuals with long COVID compared to their matched controls were further elevated 12 months before long COVID diagnosis, then decreased to a near pre-pandemic level for most investigated diagnoses (Figure 2). A notable exception was asthma and dyspnea, which remained at a doubled OR 6 months post-long COVID diagnosis. Thus, it appears that long COVID may trigger the onset of asthma. The phenomenon that viral infections may triggers late onset asthma in susceptible individuals is well established, particularly in women. Here we observed the same relative increase in asthma also in men. The nature of this link has not been sufficiently explored [9].

Previous experiences from the H1N1 influenza outbreak, indicated that patients with asthma were at a higher risk for severe illness, including hospitalization. However, early findings suggested that a diagnosis of asthma alone did not increase the risk of SARS-CoV-2 infection or the severity of COVID-19 outcomes. There is growing evidence that different asthma phenotypes play a significant role in assessing the risk of SARS-CoV-2 infection and severity of illness. Studies suggest that Th2-high inflammation may actually lower the likelihood of infection and severe outcomes, while Th2-low asthma may be linked to an elevated risk (22-24). Our findings that ICD-10 codes related to non-allergic asthma are a risk factor for long COVID development support these findings also for the SARS-CoV-2 virus. Our study further shows that asthma is associated with higher risk of persistent symptoms leading to long COVID. This suggests that while asthma may not be a significant driver of acute COVID-19 severity, it poses an increased risk for the development of long-term complications related to long COVID.

Our findings suggest that both female sex and asthma are associated with an increased risk of developing long COVID. The fact that female sex is a well-established risk factor for asthma raises the question of whether female sex is an independent risk factor for long COVID, or if the observed association is partially mediated by the higher prevalence of asthma among women.

Stratified analyses revealed that both men and women with asthma have a significantly higher risk of long COVID compared to controls without asthma. Interestingly, additive risk analyses demonstrate a slightly greater risk difference among women (2.37%) than men (1.36%), suggesting that asthma and female sex may have a synergistic impact in relation to long COVID. Moreover, based on an estimated odds ratio (OR) of 2.24 for female sex as a risk factor for long COVID, this suggests that being female independently contributes to the risk of developing long COVID, even when considering the potential influence of asthma and other confounding factors.

The large population-based cohort and comprehensive healthcare data presented here offer significant strengths, and the matching of cases and controls based on age, gender and socioeconomical area minimizes potential confounding bias. However, our study has limitations that warrant consideration. We used a retrospective study design, yet all diagnoses were registered prospectively in the register we used, limiting information bias regarding the ascertainment of diagnoses. In addition, long COVID was a newly established diagnosis at the time, leading to an underdiagnosis due to a lag in clinical practice, an effect that may have been further accentuated by the general constraints on the healthcare apparatus during the on-going pandemic. As such, it cannot be excluded that many subjects from the control group may have had undiagnosed long COVID. This would bias results toward the null, rather than an overestimate.

An important aspect to consider regarding asthma diagnosis is the impact the pandemic had on the availability of care of this patient group. Access to dynamic spirometry which is essential both for diagnosis and long-term asthma management was limited [29]. The COVID-19 pandemic has had a negative impact on asthma care, with about 25% fewer registrations of asthma and COPD patients in the Swedish Respiratory Registry in 2020 compared to 2019. This suggests that many patients did not undergo the necessary spirometry tests, likely due to the reallocation of healthcare resources and the shift to digital follow-up visits. The risks to healthcare workers related to aerosol formation during spirometry may also have limited the number of spirometry tests performed, particularly for more severe patients. According to the Respiratory Registry, individuals with asthma and COPD did not receive the expected level of evaluations and follow-ups, resulting in fewer asthma diagnoses than usual. The reduced use of spirometry may also be attributed to COVID-19-related guidelines advising against such tests [30]. The global pandemic caused major disruptions in healthcare systems, altering guidelines, medical procedures, and patient interactions. These changes likely contributed to delays in medical assessments and diagnoses, leading to underdiagnosis. The reduced availability of in-person visits, particularly in primary care, may have delayed new diagnoses, and this trend could have extended to other symptoms recorded later as healthcare providers adapted to new protocols. It is also important to recognize that this analysis is based on electronic health records, which often focus on specific reasons for visits and may not fully reflect a patient’s overall health status.

## Conclusions

Our study shows that women and individuals with pre-existing respiratory conditions, including asthma, had a higher risk of developing long COVID after primary SARS-CoV-2 infection during the first two pandemic waves in the Stockholm Region, representing the wild type and alfa strains. Future research to explore the mechanisms linking pre-existing respiratory diseases to long COVID is needed to identify specific sub-populations of long COVID for better clinical management and targeted interventions.

## Author contributions

Study design and conception: PL, ACC, GL, SL, ÅMW

Data acquisition: ACC

Quality control of data and algorithms: GL and ACC

Data analysis and interpretation: all authors

Statistical analysis and data presentation: GL, PL, ÅMW, IK

Manuscript preparation: PL, ACC, ÅMW

Funding and infrastructure: ÅMW, ACC

Manuscript editing: all authors

Manuscript review: all authors

## List of abbreviations

Long COVID
VAL: Stockholm Regional Health Care Data Warehouse
ICD-10: International Classification of Diseases, 10th edition;
OR: odds ratio
CI: confidence interval

## Supporting information

VAL

## Data Availability

All data produced in the present work are contained in the manuscript

