## Supplementary material for "Association of pre-pandemic respiratory system diseases with Long COVID: a population-based case-control study": VAL

**Supplementary Table 1. ICD-10 diagnoses extracted from VAL database**

| Code | Diagnosis |
|---|---|
| **J06.9** | Acute upper respiratory tract infection |
| **J20.9** | Acute bronchitis |
| **J40.9** | Bronchitis unspecified as acute or chronic |
| **J45.1** | Non-allergic asthma |
| **J45.8** | Mixed asthma |
| **J45.9** | Asthma, unspecified |
| **J46.9** | Acute severe asthma |
| **R05** | Cough |
| **R06.0** | Dyspnea |
| **R06.8** | Other and unspecified respiratory disorders |

**Supplementary Table 2.** Age and group sizes based on gender

| Gender | Group | N | Age* |
|---|---|---|---|
| **Female** | Case | 3 862 | 47 (39-55) |
| **Male** | Case | 1 727 | 48 (38-57) |
| **Female** | Control | 32 151 | 47 (39-55) |
| **Male** | Control | 15 410 | 47 (38-57) |

** Age expressed as median (IQR)*

**Supplementary Table 3.** Frequency of studied diagnoses in females by time period. Female included Cases n=3 862, Controls n=32 151.

| Diagnoses | Pre-pandemic (year 2019) | | 12 months pre- long COVID diagnosis | | 6 months post-long COVID diagnosis* | |
|---|---|---|---|---|---|---|
| | Case | Control | Case | Control | Case | Control |
| Acute upper respiratory tract infection J06.9 | 461 (11.9%) | 1852 (5.60%) | 349 (9.04%) | 535 (1.66%) | 79 (2.05%) | 212 (0.66%) |
| Asthma J45.1, J45.8, J45.9, J46.9 | 193 (5.00%) | 845 (2.63%) | 306 (7.92%) | 722 (2.25%) | 174 (4.51%) | 407 (1.27%) |
| Cough R05 | 178 (4.61%) | 849 (2.96%) | 334 (8.65%) | 497 (1.55%) | 106 (2.74%) | 284 (0.88%) |
| Acute bronchitis J20.9, J40.9 | 96 (2.49%) | 376 (1.17%) | 63 (1.63%) | 97 (0.30%) | 17 (0.44%) | 61 (0.19%) |
| Dyspnea R06.0 | 62 (1.61%) | 268 (0.83%) | 621 (16.1%) | 409 (1.27%) | 258 (6.68%) | 195 (0.61%) |
| Other and unspecified respiratory disorders R06.8 | 6 (0.16%) | 15 (0.05%) | 29 (0.75%) | 13 (0.04%) | 7 (0.18%) | 6 (0.02%) |

* *The 6-month time point was truncated for some individuals due to data access. Please see methods.*

**Supplementary Table 4.** Frequency of studied diagnoses in males by time period. Male included Cases n=1 727, Controls n=15 410.

| Diagnoses | Pre-pandemic (year 2019) | | 12 months pre- long COVID diagnosis | | 6 months post-long COVID diagnosis* | |
|---|---|---|---|---|---|---|
| | Case | Control | Case | Control | Case | Control |
| Acute upper respiratory tract infection J06.9 | 126 (7.30%) | 474 (3.08%) | 103 (5.96%) | 146 (0.95%) | 29 (1.68%) | 62 (0.40%) |
| Asthma J45.1, J45.8, J45.9, J46.9 | 55 (3.18%) | 281 (1.82%) | 105 (6.08%) | 234 (1.52%) | 62 (3.59%) | 126 (0.82%) |
| Cough R05 | 54 (3.36%) | 294 (1.91%) | 154 (8.92%) | 189 (1.23%) | 49 (2.84%) | 96 (0.62%) |
| Acute bronchitis J20.9, J40.9 | 33 (1.91%) | 109 (0.71%) | 28 (1.62%) | 32 (0.21%) | 5 (0.29%) | 18 (0.12%) |
| Dyspnea R06.0 | 19 (1.10%) | 89 (0.58%) | 244 (14.1%) | 161 (1.04%) | 100 (5.79%) | 73 (0.47%) |
| Other and unspecified respiratory disorders R06.8 | 0 (0.00%) | 4 (0.03%) | 4 (0.23%) | 11 (0.07%) | 4 (0.23%) | 4 (0.03%) |

* *The 6-month time point was truncated for some individuals due to data access. Please see methods.*
